# Effect of Women’s View Toward Their Rights on Receiving Better Antenatal Care in Bangladesh

**DOI:** 10.1101/2023.07.03.23292132

**Authors:** Ramisa Morshed

## Abstract

**Background:** In a country, many girls may forcefully be married off at an early age where they might have no hand in it. But the thought and view they carry throughout their lifetime can be resembled in their children’s lives too and can proclaim plenty of the country’s problems, especially receiving improved antenatal care (ANC). A woman or a prospective mother’s view of her husband beating her tells a lot about a country’s view, and a woman’s preferred age of marrying can also exhibit her attitude toward her rights and goals. However, limited research has focused on understanding women’s perspectives on their rights and their relation with ANC quality in Bangladesh. The findings of this study will contribute to a deeper understanding of women’s perspectives on their rights and the barriers they face in accessing quality ANC.

**Objective:** This study aims to investigate the view of Bangladeshi women toward their own rights and the impact on their access to improved ANC.

**Method:** To address this gap, a binary logistic regression model was employed by analyzing in IBM SPSS Statistics 25 using Bangladesh Demographic and Health Survey (BDHS) 2017-18 data. The study recruited a diverse sample of women from various social backgrounds, beliefs, and geographical locations in Bangladesh.

**Result:** It was spotted that a small percentage (6.6%) of women in Bangladesh were under proper ANC coverage. The women who didn’t justify domestic violence were 1.231 times more likely to be under ANC coverage in Bangladesh than those who justified domestic violence in the adjusted model. Moreover, women with no education were .048 times less likely to be under ANC coverage than women with higher education. Women who were not exposed to media were .099 times less likely to be under ANC coverage than those who were exposed to media. Unfortunately, almost one-fifth of all Bangladeshi women (19.6%) justified being

**Conclusion:** Women’s entitlements enable them to actively engage in their own healthcare programs, leading to improved health outcomes for both mother and child. Through targeted awareness campaigns, educational initiatives, and policy advocacy, beaten by their husbands.

Bangladesh can create an environment where women are aware of their rights and have access to quality ANC services. By ensuring their rights are respected, a society that values reproductive health and the well-being of women and children can strive.

## Introduction

Women’s involvement in understanding and establishing their rights (here it is resembling if the mothers can say no to domestic violence and if they discourage an early marriage) in Bangladesh may not at all times be prioritized. In many societies, women are often restricted from understanding their worth regarding their own well-being and health, with their rights being dictated by their husbands. Consequently, this can result in inadequate or inappropriate care for women during pregnancy, negatively impacting their health and the well-being of their babies. Anyway, it remains uncertain whether women’s view about their own rights directly relates to improved ANC in Bangladesh. This topic holds noticeable importance as ANC plays a critical role in ensuring a healthy pregnancy and reducing maternal and infant mortality rates. According to a report in 2015, approximately 300,000 women worldwide died annually due to birth giving related complications, with a majority of these deaths being preventable [17]. In 2020, the number of global maternal mortalities came to 2,80,000 [16].

ANC encompasses various aspects such as regular check-ups, tests, and counseling, all aimed at promoting a healthy pregnancy and delivery. Factors such as the timing of the first ANC visit, the frequency of visits, and seeking care from professionals greatly influence the effectiveness of maternal health interventions during the antenatal period. According to “the WHO FANC model”, it is required that the mother’s 1st ANC visit must be within the first 12 weeks of gestation [18]. It is also mandatory that under usual circumstances, a prospective mother should have a minimum of four ANC visits [19]. Not receiving a proper ANC at least four times might diminish the likelihood of receiving a worthwhile maternal health intervention during the antenatal period [20,21]. Despite an increase in ANC coverage in Bangladesh, the country still ranked among the top ten contributors to global maternal mortality rates in 2015. Although there has been some improvement in recent years, it remains insufficient compared to other developed countries [22]. Therefore, it is crucial to identify the impact of women’s upholding their rights related to marriage on ANC utilization.

Previous studies have shown that receiving ANC from skilled providers reduces the risk of complications and adverse pregnancy outcomes; such as-intrauterine growth retardation, stillbirths, preterm births, fetal abnormalities, etc [7-15]. However, in Bangladesh, only a small percentage of women received their first ANC within the recommended timeframe, highlighting the need for improved access to quality care. In 2016, it is manifested that just about one-fifth (21%) of Bangladeshi women received their first ANC within 3 months of gestation from any health service provider; more agitatedly only 14% of those women received ANC from a skilled service provider [23].

Women’s attitude to their rights can significantly influence the standard of care they receive during pregnancy and childbirth. When women have the mentality or attitude to say no to violence and early marriage, they might be more likely to seek care early in their pregnancy, attend all recommended appointments, and follow healthcare providers’ advice; which was investigated through this analysis. This, in turn, can lead to better health outcomes for both mothers and babies, as well as a more positive childbirth experience and improved health for women. In Rwanda, there was a significant negative relationship between physical intimate partner violence and both early ANC and sufficient ANC [24]. In India, the results of a study revealed that women who married at <18 years were significantly less likely to use maternal health care services than those married at ≥18 years even after accounting for socio-economic and demographic characteristics of women [25]. Once more, a research about child marriage in south Asian countries showed that the common factors resulting from child marriage were reproductive and maternal factors such as reduced utilization of ANC services, fewer institutional deliveries, and the absence of a skilled birth attendant during delivery [26]. Again, in Bangladesh, domestic violence was significantly associated with the lower utilization of four or more ANC and health-center-based delivery care [27]. But no research showed the urgency of the overall ANC coverage that includes the required times to visit for ANC, the first ANC visit time, and going to qualified centers or doctors; with the effect of women’s attitude to violence and their preferred age for marriage.

The primary goal of raising awareness among women about their marriage rights and improving access to quality ANC is to ensure their overall well-being during the crucial stages of pregnancy. The importance of women’s rights and equitable healthcare provision during pregnancy cannot be overstated for this country, as it directly influences maternal and child health outcomes. Educating women about their rights within the institution of marriage, such as the right to consent, equips women to make informed choices regarding their reproductive health, including seeking timely and appropriate ANC. Simultaneously, enhancing access to better ANC services is essential for promoting positive maternal and child health outcomes. By providing comprehensive and evidence-based care during pregnancy, ANC plays a vital role in detecting and managing potential risks, ensuring healthy pregnancies, and reducing maternal and antenatal mortality rates.

However, there is a lack of research specifically examining the impact of women’s view to say no to the violation of their rights on receiving quality ANC in Bangladesh. Further investigation is obligatory to understand the important relationship between women’s view about their rights and ANC utilization in this country. It is essential to recognize the significance of enlightening women and ensuring the good for their own health and the health of their families. By promoting women’s attitude toward their rights and saying no, Bangladesh can endeavor toward providing adequate ANC and improving maternal and antenatal health outcomes in Bangladesh.

## Data and Variables

The data from the Bangladesh Demographic and Health Survey (BDHS) of the 2017-18 year has been used in this analysis. It contains the data of Bangladeshi married women who ever had a child. Finally, the analysis was done using the data of 10582 respondents. The process of ending up to 10582 respondents after data cleaning is discussed in the Methodology section.

The independent variables that were used to predict the dependent variable “ANC Coverage” were “Woman’s justification of being beaten by husband” and “Preferred age for marriage”. The covariates were: Highest educational level, Religion, Type of place of residence, and Exposure to media. It is to mention that covariates were selected seeing the highest Cox and Snell R square doing backward elimination.

### Dependent Variable

- **ANC Coverage:** ANC coverage was calculated based on three factors. If the woman goes for her first ANC visit within the first three months of conceiving, had at least four ANC visits during her whole pregnancy period, and went to qualified centers/doctors for ANC visits were assumed to be under ANC coverage. Here the qualified centers or doctors were all the govt. medical centers and hospitals, private medical centers and hospitals, and qualified doctors. The clinics in local area or community or ANC at home were not assumed as quality ANC. Specific factors behind being under ANC coverage were analyzed in this research.

Independent Variables

### Women’s view to their rights

In this analysis, women’s view to their rights resembles if the mothers can say no to domestic violence and if they discourage early marriage. So this attitude was manifested into two independent variables.

1. **Women’s justification of being beaten by husband:** In this analysis, the factors that might affect being under ANC coverage were related to women’s view to their rights. That means how married women in Bangladesh who ever had a child behave or think about their own good and self-respect. The women who think being beaten by husbands for any reason is okay and justified are not carrying the same attitude that the other women do, who think beating is not justified for any reason. The reasons were if women go out without telling their husbands, neglect their children, argue with husband, refuse to have sex with their partners, and burn the food. Women who think being beaten for any of these reasons is okay justify beating.
2. **Preferred age for marriage:** In the logistic regression and further calculation, the quantitative variable “Preferred age for marriage” was used. In percentage analysis (table 1), the preferred age for marriage was grouped into 3 classes for representing the percentage easily as it was impossible without grouping the quantitative variable like age. But the quantitative raw variable was used in the probit model. In Bangladesh, the minimum age of marriage for girls is 18 years [1]. In the following dataset, it was seen that many women prefer marrying at the minimum age which is 18, and even before 18. The preferred age of marriage according to the individuals also indicates their view to their marriage rights and planning for their future and own selves.

**Table 1:** Percentages of married women under every category of variables who ever had a child

| Variables | Percentages of respondents |
| --- | --- |
| **ANC coverage** |  |
| Under quality ANC coverage | 6.6% |
| Not under quality ANC coverage | 93.4% |
| **Highest Educational Level** |  |
| No education | 15.9% |
| Primary | 31.5% |
| Secondary | 38.6% |
| Higher | 14% |
| **Type of place of residence** |  |
| Urban | 36.6% |
| Rural | 63.4% |
| **Religion** |  |
| Muslim | 90% |
| Non-Muslim | 10% |
| **Exposure to media** |  |
| Connected to any news media at least once a week | 55.5% |
| Not connected to any news media in a week | 44.5% |
| **Preferred age for marriage** |  |
| Prefers marrying before 18 | 2.6% |
| Prefers marrying exactly at 18 | 74.6% |
| Prefers marrying after 18 | 22.8% |
| **Woman's justification of being beaten by husband** |  |
| Justifies beating for at least one reason | 19.6% |
| Doesn't justify beating for any reason | 80.4% |

**Table 2:** Independent variables with p-value attained from Chi-square test of association

| <u>Variables</u> | <u>p-value</u> |
| --- | --- |
| Woman's justification of being beaten by husband | <.001 |
| Preferred age for marriage | <.001 |
| Exposure to media | <.001 |
| Highest educational level | <.001 |
| Type of place of residence | <.001 |
| Religion | .336 |

**Table 3:** Binomial Logistic Regression, all ever-married women who ever had a child

|  |  | **Model 1 (Unadjusted)** |  | **Model 2 (Adjusted)** |  |
| --- | --- | --- | --- | --- | --- |
|  |  | Odds Ratio | S.E. | Odds Ratio | S.E. |
| Woman's justification of being beaten by husband | **Yes (RC)** |  |  |  |  |
|  | No | 1.537*** | .117 | 1.231* | .120 |
| Preferred age for marriage |  | 1.083*** | .028 | .986 | .030 |
| Exposure to media | **Yes (RC)** |  |  |  |  |
|  | No |  |  | .658*** | .099 |
| Highest Educational Level | **Higher (RC)** |  |  |  |  |
|  | No education |  |  | .048*** | .302 |
|  | Primary |  |  | .212*** | .148 |
|  | Secondary |  |  | .527*** | .125 |
| Type of place of residence | **Rural (RC)** |  |  |  |  |
|  | Urban |  |  | 1.096 | .091 |
| Religion | **Muslim (RC)** |  |  |  |  |
|  | Non-Muslim |  |  | .756 | .175 |
| Constant |  |  |  | .208** | .612 |

In this analysis, a single star (\*) means a significance level of 10%, a double star (\*\*) means a significance level of 5%, and a triple star (\*\*\*) means a significance level of 1%. Here the highest acceptable level of significance is 10%.

### Covariates

1. Highest educational level: Four educational levels were labeled in the highest education variable to check if there were any significant changes in ANC coverage with different educational levels. It was used to adjust the model.
2. Type of place of residence: This demographic variable was also used as a covariate and was used to adjust the model in logistic regression. It can show whether rural or urban people were more likely to be under ANC coverage.
3. Religion: The religion variable from BDHS data was again divided into two categories-Muslims and Non-Muslims. This variable also shows the likelihood of being under ANC coverage among two categories of religion and was mainly employed to adjust the model.
4. Exposure to Media: Respondents who were connected to any single news media at least once a week were assumed to be exposed to media; and those who don’t watch or listen to any type of news media even once a week were not exposed to media. This created variable was also used to adjust the model.

It was seen that these socio-demographic variables put an impact in adjusting the model where the dependent variable was ANC coverage [2]. Though it might vary regarding the data, social changes at different times in a country, and the goal of a study.

## Methodology

The analysis was performed using IBM SPSS Statistics 25 with the data of 10582 respondents taken from BDHS 2017-18 data. In this research, it was to see if the factors that determine women’s view toward their own rights put a significant impact on receiving better ANC. To understand women’s view toward their own rights, their preferred age to get married and their justification status of being beaten by their husbands were used here.

The missing data were deleted from the study except for the missing data from the ANC-related variables such as where they received their ANC from. As the whole dataset contains the data of mothers, they all must have gone through the pregnancy period and if they didn’t answer the quality ANC-related questions but answered all other queries, it was more likely that they actually didn’t get a proper ANC. So deleting the missing data would create a bias here. The women who answered the other questions but ignored the answer to where they received the ANC from were assumed not to be under ANC coverage.

ANC coverage is a dichotomous variable. Women who had their first ANC visit within the first three months of conceiving, had a total of 4 ANC visits minimum, and went to qualified centers or doctors were assumed to be under better ANC coverage. The independent variable Woman’s justification of being beaten by husband was calculated like-beating is justified or coded 1 if that woman thought that beating for any of the following reasons is justified (discussed in the variables section). Thus religion groups and Media exposure were also coded into two categories as discussed before.

A chi-square test was conducted to see the association between the ANC coverage and the other factors individually. Then the binary logistic regression was run and it gave an unadjusted and an adjusted model that was adjusted with covariates. The odds ratios from logistic regression models showed how the likelihood of occurring the event (being under ANC coverage) was for each category of the categorical regressors and a quantitative regressor. The selected regressors for logistic regression didn’t have a strong correlation with each other.

An odds ratio (OR) is a measure of the association between an exposure and an outcome. The OR represents the odds that an outcome will occur given a particular exposure, compared to the odds of the outcome occurring in the absence of that exposure [3]. The logistic regression equation is written as log (P/(1-P)) = Beta* X + Intercept; where log (P/(1-P)) is called the log odds or logit. P is the probability of having the outcome and P/(1-P) is the odds of the outcome. Thus, the exp(B) shows the odds of happening an event for that category compared to the reference category.

## Result

### Descriptive Statistics

In Table 1, the percentage analysis is shown for every variable used. As previously said in the introduction, women must have at least one ANC visit within the first three months of conceiving and they must receive at least four ANC visits within their whole pregnancy period. These two things and going to quality professionals for ANC were combined into the final variable ANC coverage.

Surprisingly, it can be seen that only 6.6% said yes to all these three factors (going for the first ANC visit within the first three months, having a total of 4 ANC visits, and going to quality professionals for ANC). So the rest 93.4% were not under the ANC coverage in Bangladesh. More than half of the married females’ sample with children have completed at least their secondary education. The majority of the sample were Muslims and were from Rural areas. Shockingly, 19.6% of the whole female sample of Bangladesh justify being beaten by their husbands.

The **bar diagrams** shown below reveal the **percentages of women who received better ANC** under every category of each variable.

**Fig 1:**
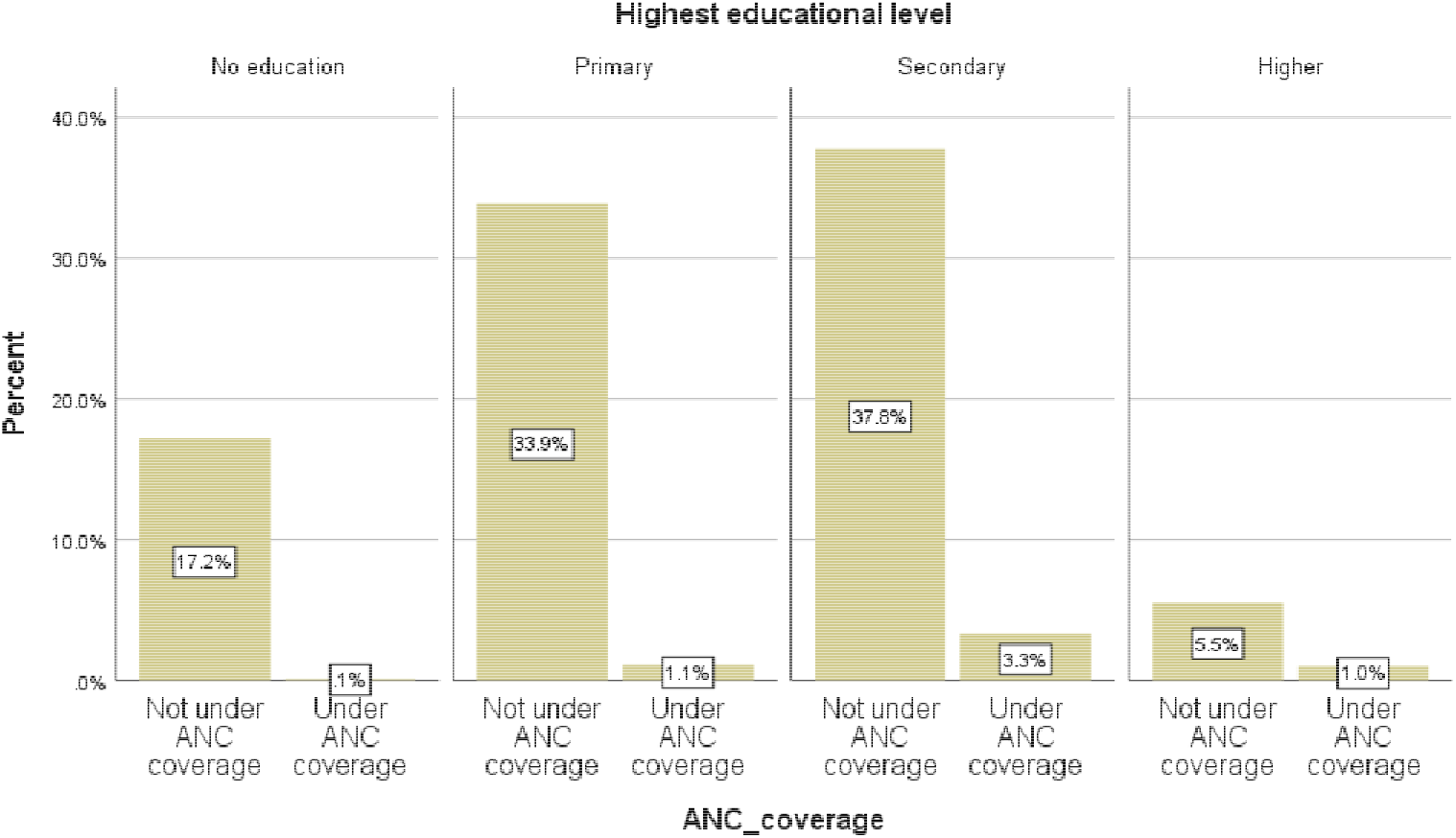
Bar diagram showing percentage of women who were in different educational levels from two ANC coverage status

**Fig 2:**
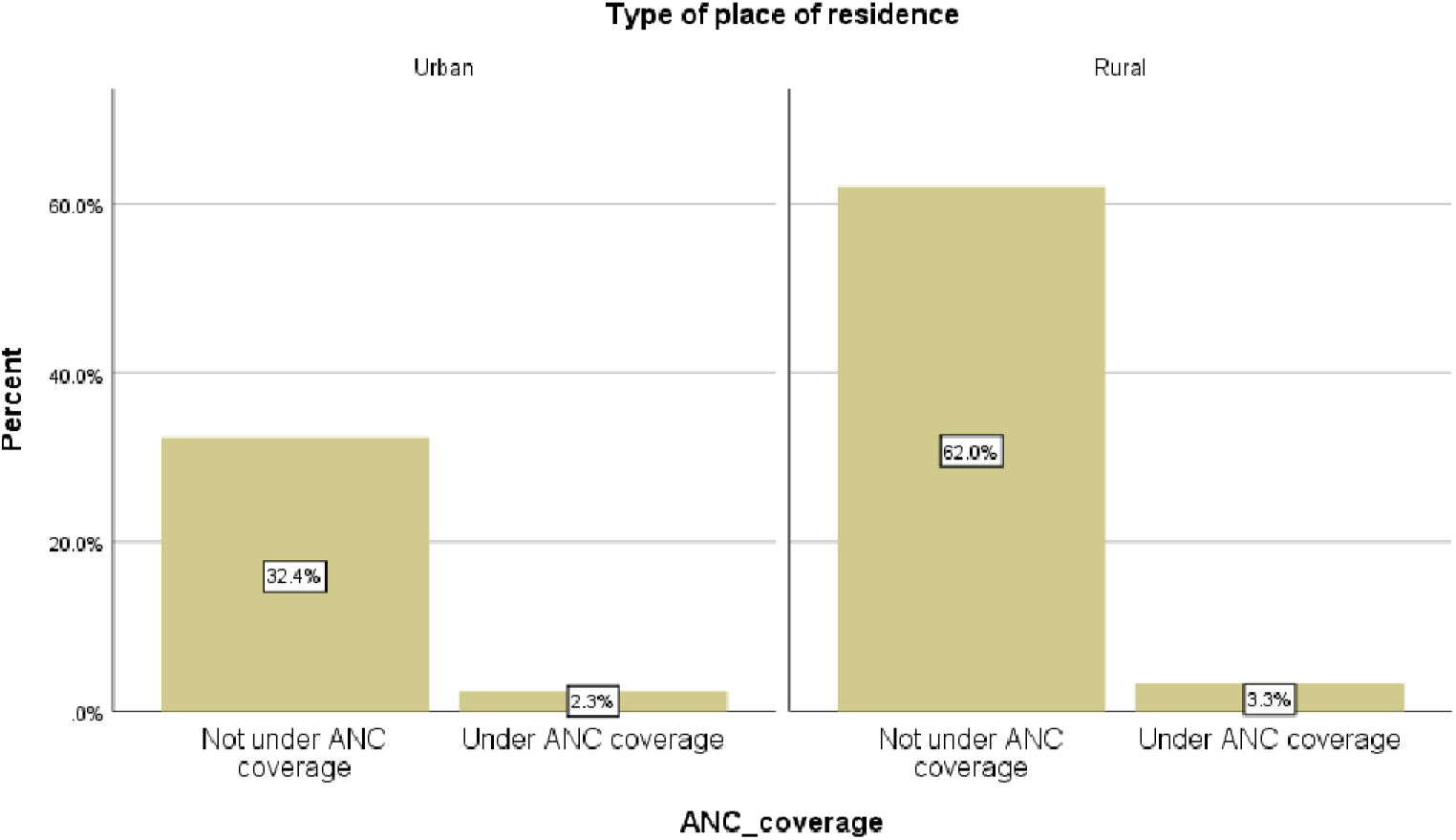
Bar diagram showing percentage of women who were from different types of residence from two ANC coverage status

**Fig 3:**
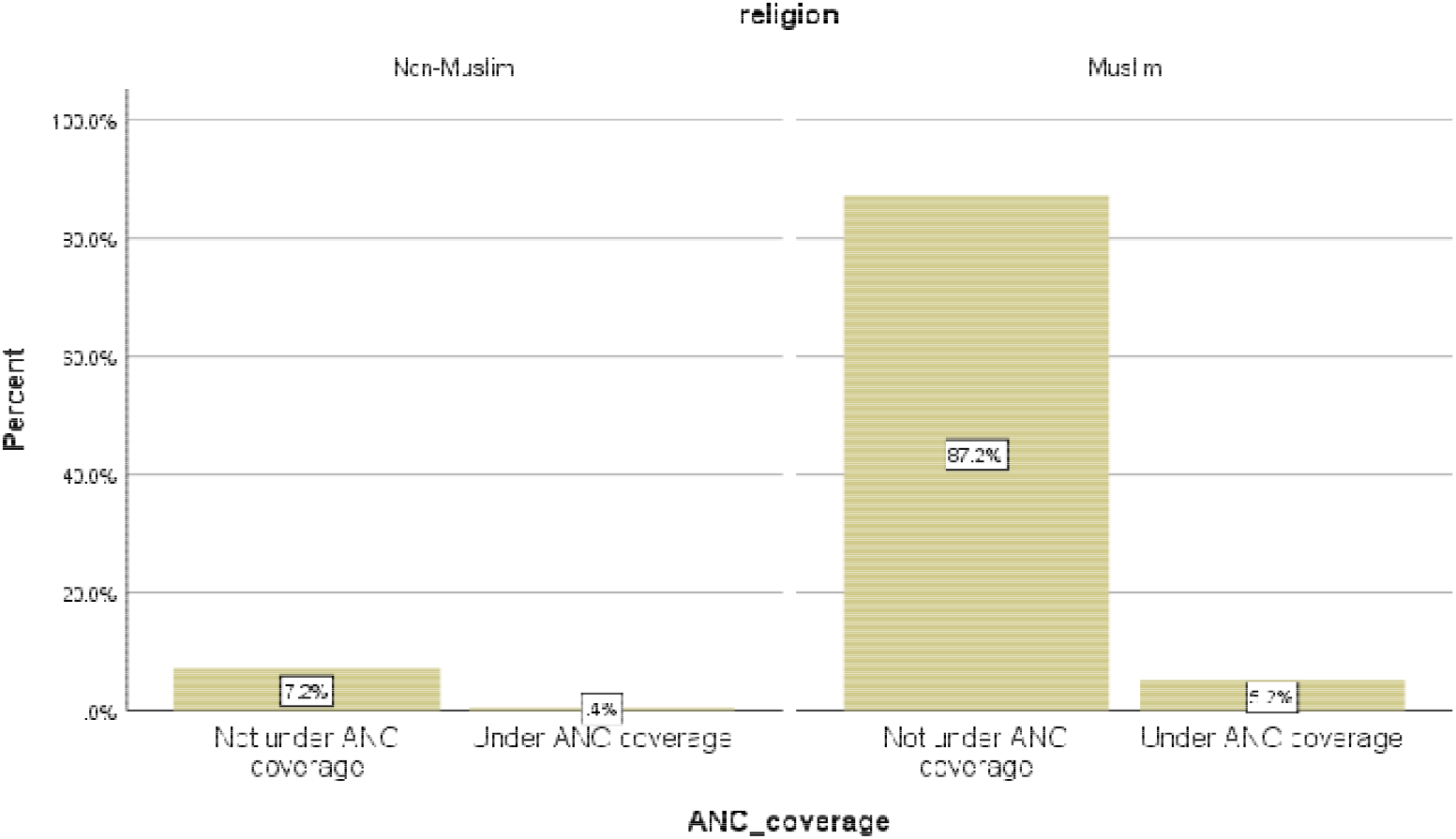
Bar diagram showing percentage of women who were from different religion (Muslims and Non-Muslims) from two ANC coverage status

**Fig 4:**
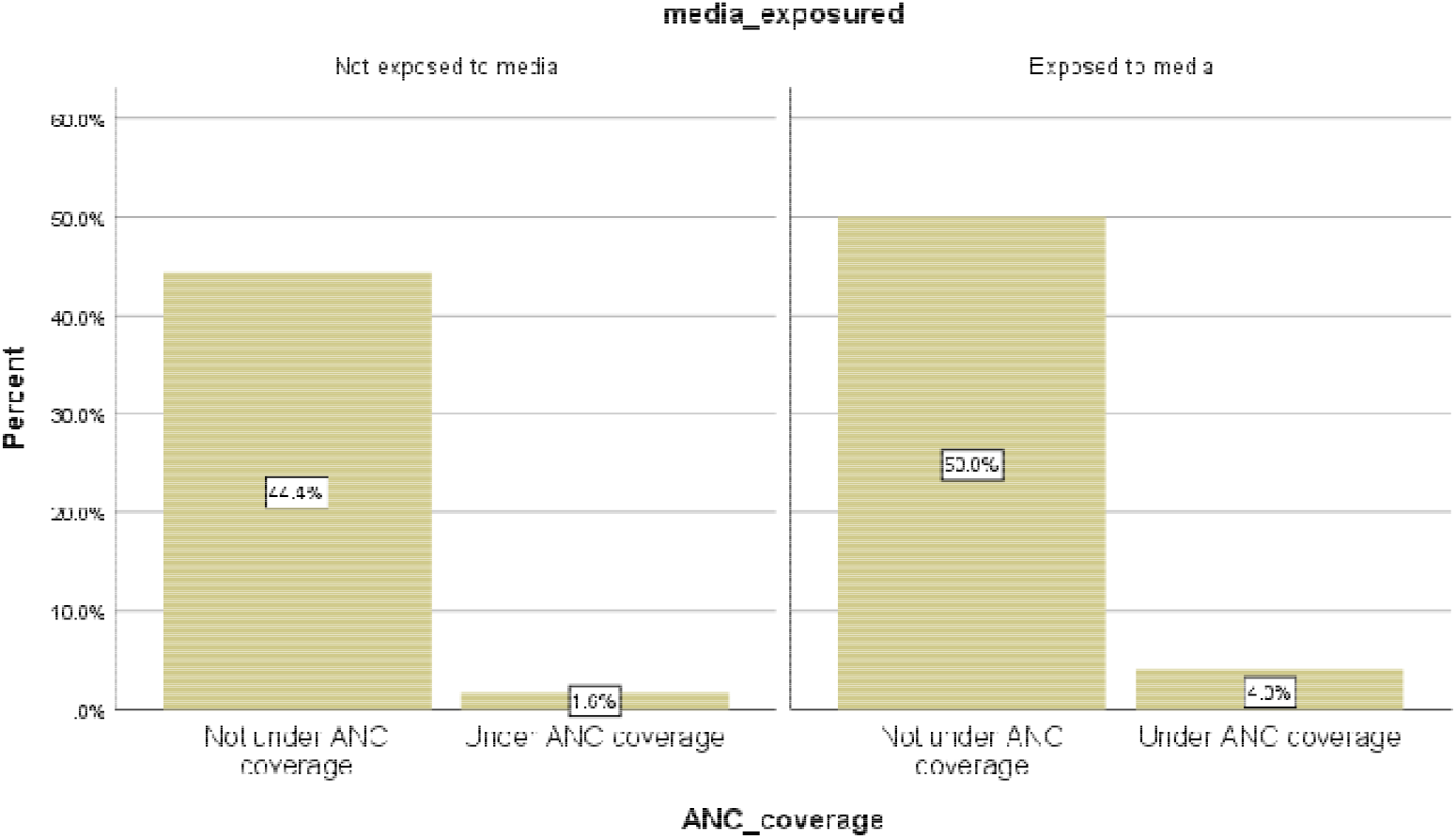
Bar diagram showing percentage of women who were from different levels of media exposure from two ANC coverage status

**Fig 5:**
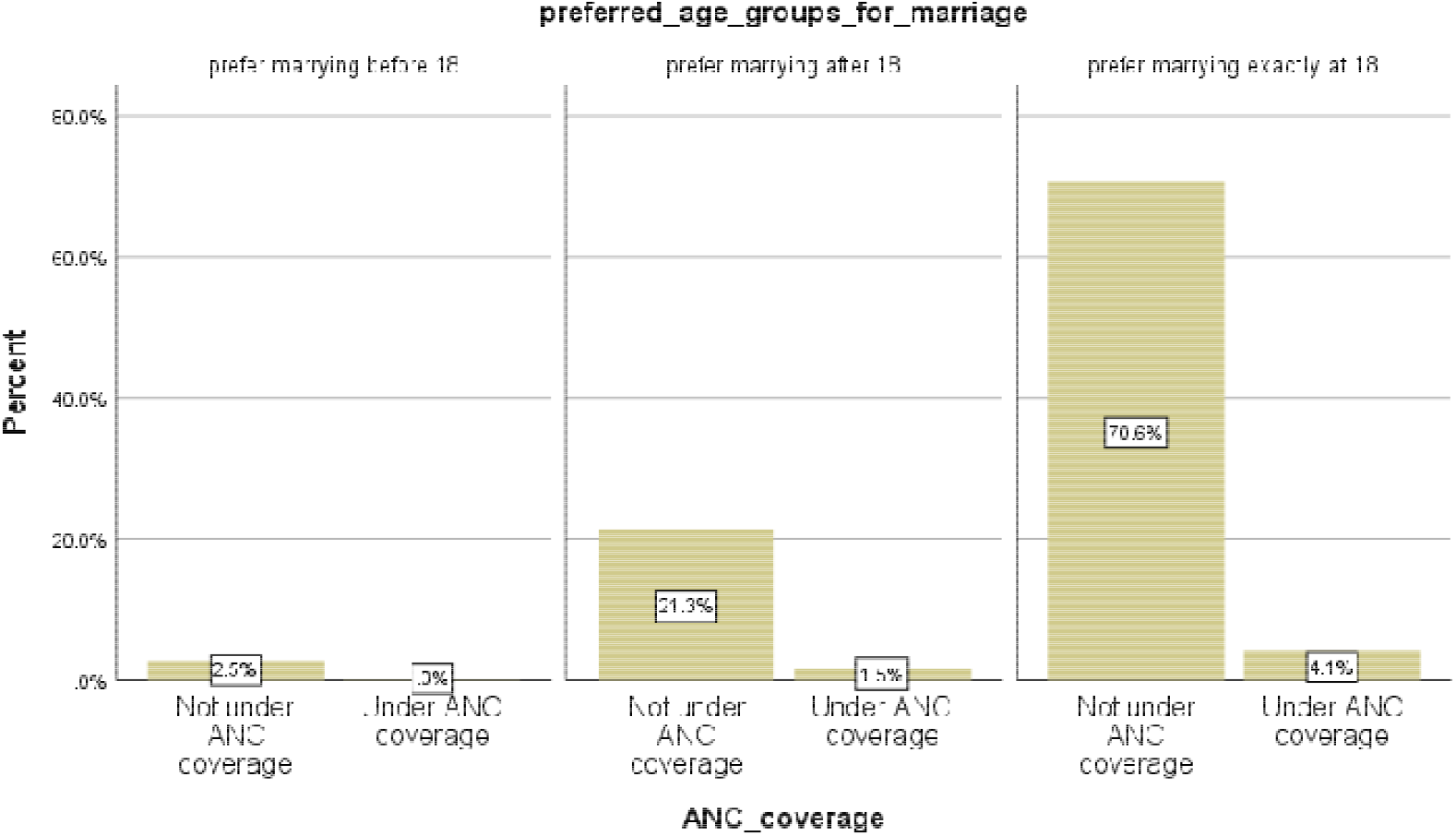
Bar diagram showing percentage of women preferring different marriage age groups from two ANC coverage status

**Fig 6:**
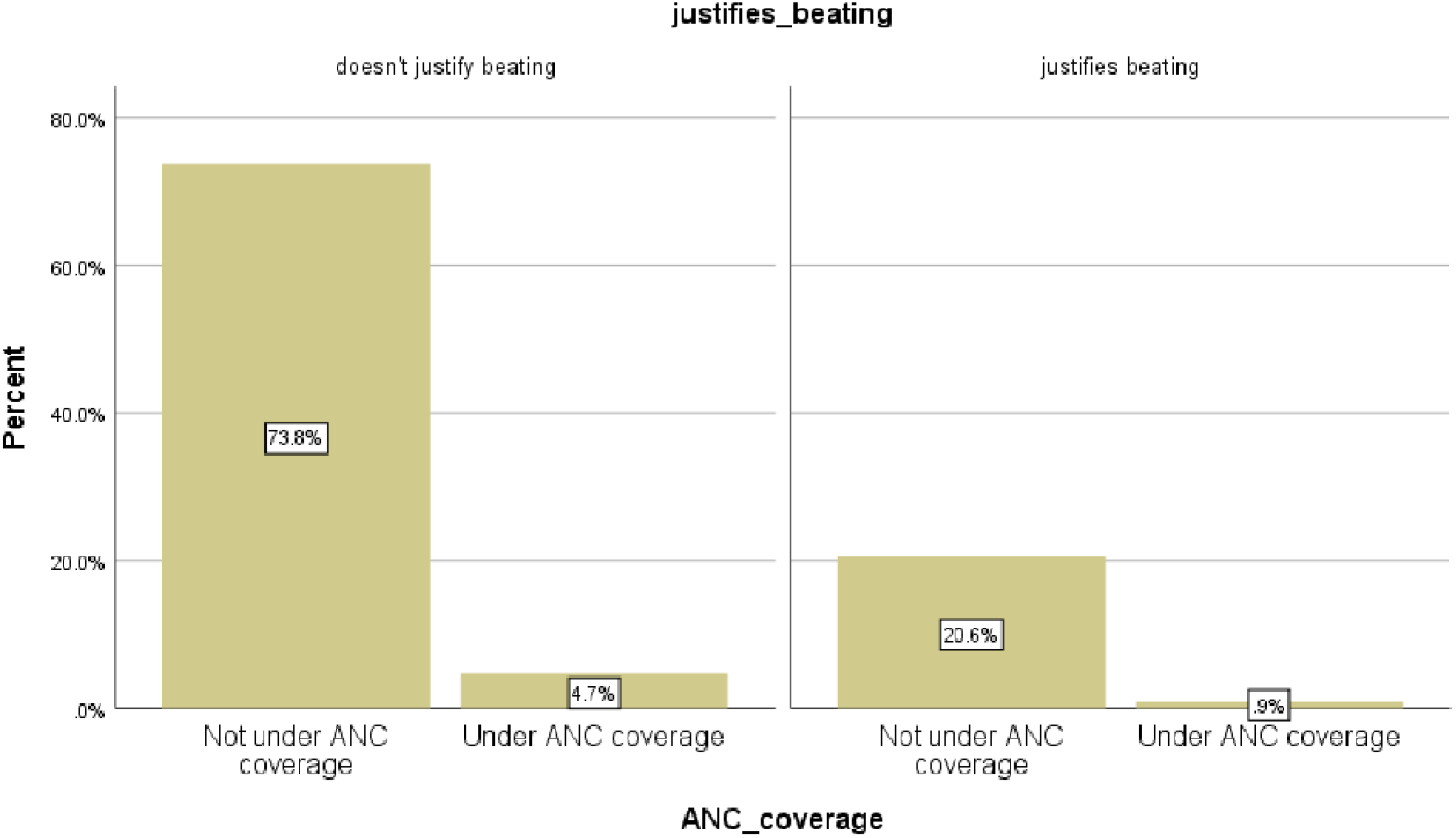
Bar diagram showing percentage of women who justify beating from husbands from two ANC coverage status

A chi-square test was conducted with the dependent variable to all the independent variables and covariates that were applied in the probit model.

All the independent variables and covariates had a significant association with ANC coverage except the Religion groups. A probit model shows the likelihood of occurring an event that is being under ANC coverage here.

### Results of the Probit Model

A binary logistic model is a probit model where an outcome variable can be of two types only. Here either the outcome can be under ANC coverage or not under ANC coverage. The exp(B) or odds ratio tells how the likelihood of occurring an event is (here, it is being under ANC coverage) for each category of the regressors compared to the reference category. The **reference category** is a category under each categorical variable with which the likelihood of occurring an event for other categories is compared.

## Discussion

It was seen from the descriptive statistics that the number of respondents (70.6% of the whole sample) preferring marrying exactly at 18 was huge in the category who were not under ANC coverage. Unfortunately, 37.8% of the sample and 5.5% of the sample were from secondary and higher educational levels respectively who were not under the ANC coverage. Moreover, 32.4% of the sample were from urban areas and were not into ANC coverage, whereas people tend to think that urban people are privileged enough. More astonishingly, 50% of the sample were exposed to media and were still not under the ANC coverage.

In the unadjusted probit model which is Model 1 (Wald statistic: 4459.364***, Cox and Snell R square: .002), both Woman’s justification of being beaten by husband and Preferred age for marriage came out to be significant regressors of the model. The result in the 1st model says that, the women who don’t justify being beaten by their husbands for any reason were 1.537 times more likely to receive a better ANC (being under ANC coverage) in Bangladesh than those who justify being beaten by their own husbands. Moreover, women preferring a higher age for marriage were 1.083 times more likely to receive a better ANC than those who prefer one-unit lower age for marriage.

However, in the adjusted probit model which is Model 2 (Wald statistic: 4459.364***, Cox and Snell R square: .032), Preferred age for marriage didn’t come out to be a significant factor. In that case, the odds of being under ANC coverage for women who don’t justify beating was 1.231 times higher than those who justify beating. On top of that, the highest educational level and media exposure came out to be significant covariates. It conveys that women with no education were .048 times less likely to be under ANC coverage than women with higher education. Similarly, women with a primary education were .212 times less likely and women with a secondary education were .527 times less likely to be under ANC coverage than the women with a higher education. Women who were not exposed to media that means who didn’t watch or listen to any news media for over a week were .099 times less likely to be under ANC coverage than those who were exposed to media. All these discussed results were significant.

## Conclusion

It was concluded from the analysis that women’s view to claim and hold their own rights matters in their maternal health too. Marrying at one’s preferred age (not before 18) is a woman’s right in Bangladesh [6]. Being beaten by husband is a domestic violence which is a punishable crime [4,5].

But in this analysis, it was seen what women themselves think about their own rights, and it was finally proved that it impacted significantly on ANC coverage that is closely related to a mother’s as well as a child’s health. Though the preferred age for marriage was not significant in the adjusted model but it significantly showed an effect in the unadjusted model. In addition to the beating justification, women’s education and media exposure were matters of importance too. Women need to be more educated and involved in news media for further knowledge to prevent unqualified ANC that are tremendously risky for the child and mother’s health.

Exposure to media and being highly educated can lead a woman to change her view toward violence and her own rights which may finally lead to receiving better ANC. The mentality that being beaten by one’s husband is okay for a specific reason can be extremely hazardous for a woman’s maternal health as it was deeply connected to better ANC according to this analysis. By identifying these factors that shape women’s views, healthcare providers and policymakers can develop targeted interventions to improve service delivery and enlighten women during pregnancy. Additionally, the study will shed light on potential disparities in access to ANC based on socio-demographic factors. This research is crucial for informing evidence-based policies and interventions that promote women’s view to their rights and enhance ANC services.

## Data Availability

https://dhsprogram.com/methodology/survey/survey-display-536.cfm
The data is provided by DHS from BDHS 2017-18 survey datasets

